# Natural Language Processing Can Automate Extraction of Barrett’s Esophagus Endoscopy Quality Metrics

**DOI:** 10.1101/2023.07.11.23292529

**Authors:** Ali Soroush, Courtney J. Diamond, Haley M. Zylberberg, Benjamin May, Nicholas Tatonetti, Julian A. Abrams, Chunhua Weng

## Abstract

**Objectives:** To develop an automated natural language processing (NLP) method for extracting high-fidelity Barrett’s Esophagus (BE) endoscopic surveillance and treatment data from the electronic health record (EHR).

**Methods:** Patients who underwent BE-related endoscopies between 2016 and 2020 at a single medical center were randomly assigned to a development or validation set. Those not aged 40 to 80 and those without confirmed BE were excluded. For each patient, free text pathology reports and structured procedure data were obtained. Gastroenterologists assigned ground truth labels. An NLP method leveraging MetaMap Lite generated endoscopy-level diagnosis and treatment data. Performance metrics were assessed for this data. The NLP methodology was then adapted to label key endoscopic eradication therapy (EET)-related endoscopy events and thereby facilitate calculation of patient-level pre-EET diagnosis, endotherapy time, and time to CE-IM.

**Results:** 99 patients (377 endoscopies) and 115 patients (399 endoscopies) were included in the development and validation sets respectively. When assigning high-fidelity labels to the validation set, NLP achieved high performance (recall: 0.976, precision: 0.970, accuracy: 0.985, and F1-score: 0.972). 77 patients initiated EET and underwent 554 endoscopies. Key EET-related clinical event labels had high accuracy (EET start: 0.974, CE-D: 1.00, and CE-IM: 1.00), facilitating extraction of pre-treatment diagnosis, endotherapy time, and time to CE-IM.

**Conclusions:** High-fidelity BE endoscopic surveillance and treatment data can be extracted from routine EHR data using our automated, transparent NLP method. This method produces high-level clinical datasets for clinical research and quality metric assessment.

**Study Highlights:**

1) WHAT IS KNOWN:

- Existing BE clinical data extraction methods are limited.
2) WHAT IS NEW HERE:

- An NLP pipeline for granular BE clinical data.

## INTRODUCTION

Esophageal adenocarcinoma (EAC) has been rising in incidence since the 1970s (1, 2). Despite advances in screening, surveillance, and treatment of EAC, five-year survival remains under 25% (3). Barrett’s esophagus is a premalignant precursor to EAC, characterized by a change in from normal squamous epithelium to columnar epithelium (4). US guidelines have established protocols for the endoscopic surveillance and treatment of dysplastic BE and intramucosal EAC (5–7). However, adherence to these complex guidelines has been poor (8). Quality metrics have been proposed to standardize BE-related endoscopic surveillance and treatment (9, 10), but use of this measures is limited due to challenges with efficiently abstracting high-quality clinical history and outcomes data.

Patients with BE undergo many surveillance and treatment endoscopies over the course of their lifetimes, in some cases accumulating tens of free text endoscopy and pathology reports. Billing codes have not been used to automate BE-related clinical data extraction due to inadequate code granularity and temporality (11). In the most recent US version of the International Classification of Diseases codes (ICD-10-CM), there is no code for a diagnosis of indefinite for dysplasia and, in older versions, there is only one code for all BE-related diagnoses. Moreover, ICD codes for BE cannot distinguish between prior and current diagnoses, resulting in procedure-associated diagnosis codes that represent the worst prior BE-related diagnosis, rather than the current BE-related diagnosis. Current Procedural Terminology (CPT) codes are similarly limited as they cannot distinguish among the different types of endoscopic ablative therapy (radiofrequency, cryotherapy, or argon plasma coagulation) commonly used to treat dysplastic BE and intramucosal EAC. Manual chart review remains the only viable option to date for extracting BE-related endoscopic surveillance and treatment data from clinical records. However, this approach is error-prone, not scalable, and time-consuming, limiting its use for clinical research and quality metric assessment (12–15).

Natural language processing (NLP) has improved data abstraction accuracy and efficiency for colonoscopy-related quality metrics like adenoma detection rate and bowel preparation quality, but few studies to date have created NLP tools for BE-related endoscopy (16–25). NLP pipelines for these metrics have used combinations of rule-based methods and clinical NLP tools to extract key data elements (26, 27). While most studies created NLP pipelines for clinical research, one study built a pipeline for the ongoing measurement of colonoscopy quality measures (21). A few studies included limited clinical data summarization and decision support (19, 21, 28). To date, there is a single BE-related NLP pipeline, which extracts dysplasia diagnoses from the Veterans’ Affairs electronic health records (EHR) system with high performance (29). Here we present an NLP system that automates extraction of a broader range of clinical outcomes data for both endoscopic BE surveillance and BE-related treatment, allowing automation of downstream clinical history summarization, quality metric measurement, and outcomes research.

## MATERIALS AND METHODS

### Data Source

We queried the ProVation (Minneapolis, MN, USA) clinical database to identify patients who underwent upper endoscopy between January 1, 2016 and December 31, 2020 at Columbia University Irving Medical Center (CUIMC) and had a free text procedure indication related to BE surveillance or endotherapy. ProVation is an endoscopy documentation system that captures and stores structured data in addition to transmitting a free text note to the CUIMC Clinical Data Repository (CDR). We extracted all free text pathology notes as well as structured ProVation maneuver and impression data obtained during the same period. Pathology reports were written in the Cerner CoPath (Kansas City, MO, USA) and transmitted to the CDR as free text. Free text pathology reports were ultimately extracted from both Allscripts (Chicago, IL, USA) and Epic (Verona, WI, USA) electronic health record (EHR) systems, as CUIMC transitioned from Allscripts to Epic in February 2020.

### System Development

To develop the pipeline, we randomly assigned 300 patients with a BE-related indication either to the development or validation sets. Gastroenterologists performed manual review of the EHR to determine ground truth diagnosis and endotherapy labels for each pathology report and procedure respectively. Histologic diagnosis labels included *no BE*, *no dysplasia*, *indefinite for dysplasia*, *low-grade dysplasia (LGD)*, *high-grade dysplasia (HGD)*, and *EAC*. Using these labels, a second set of simplified and clinically relevant binary diagnosis labels (presence/absence) of 1) low-grade dysplasia or worse and 2) EAC were derived. Endotherapy labels included *endoscopic mucosal resection (EMR)*, *endoscopic submucosal dissection (ESD)*, *radiofrequency ablation (RFA)*, *argon plasma coagulation (APC)*, and *cryotherapy*. One gastroenterologist reviewed records for the development set and two gastroenterologists reviewed records for the validation set. Discrepant results in the validation set were adjudicated by a third, expert gastroenterologist. We excluded patients who did not have manually confirmed diagnoses of BE (at least 1 cm of endoscopically identifiable BE and intestinal metaplasia found on esophageal biopsies) or were not 40-80 years old. We generated and troubleshooted the data processing pipeline exclusively using the development set. With each version of the pipeline, we reviewed classification errors and adjusted data processing rules as generally as possible to maximize pipeline performance. All classification errors potentially related to incorrect ground truth labels were resolved by a repeat of the initial manual review process. The validation set was used to measure performance metrics exclusively.

### Natural Language Processing

We processed pathology text using an approach combining regular expressions and MetaMapLite (30). We first excluded pathology reports lacking esophageal specimens, duplicate reports, and reports without any diagnostic information. Brushings, cytology, and surgical resections were additionally excluded. Next, we divided each pathology note into sections and excluded non-diagnostic text such as clinical history or headers. For each patient, pathology notes were linked to endoscopy impression data by date (any upper endoscopy within the 3 days preceding a pathology note). Pathology reports of externally obtained endoscopy specimens were not linked to endoscopy data, as this procedure data was not available. Endoscopies without biopsies were not linked with pathology reports. To improve medical concept recognition, we applied an expanded dictionary for BE-related terminology, incorporated additional negation logic, and removed extraneous punctuation from the pathology free text. Our system architecture is summarized in **Figure 2**.

**Figure 1.**
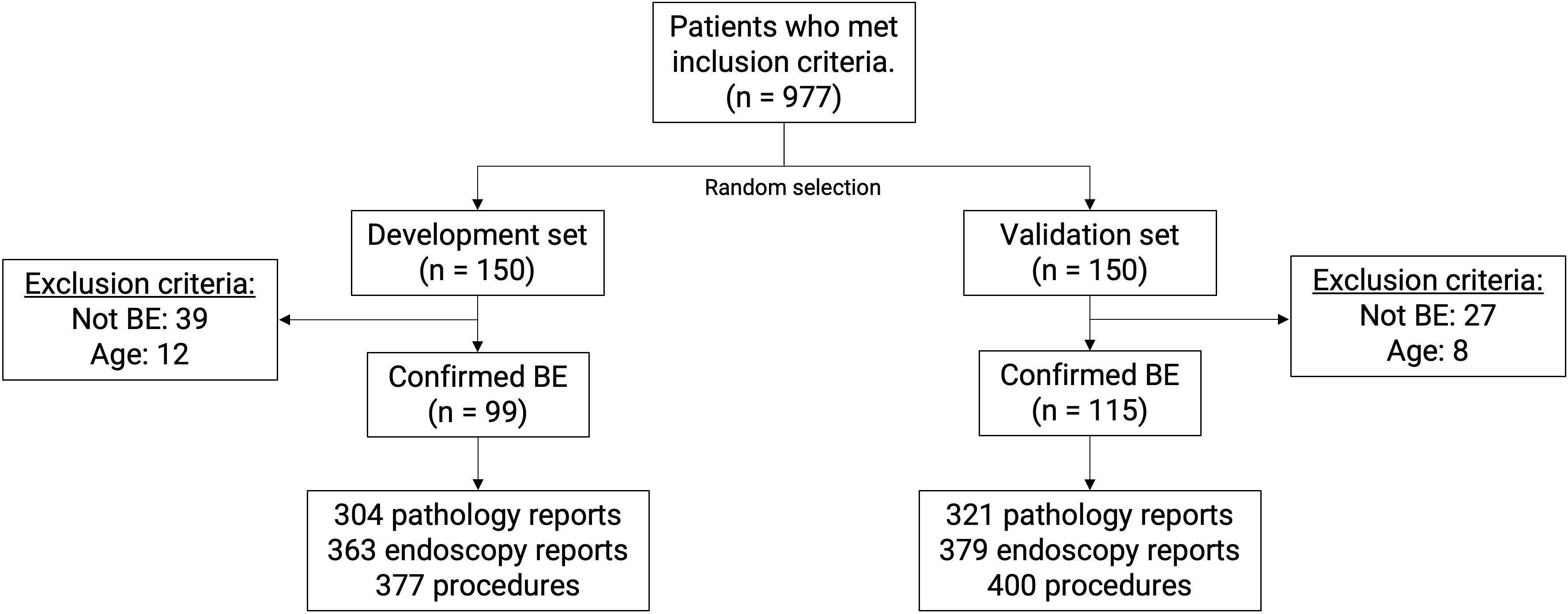
**Patient flow diagram.**

**Figure 2.**
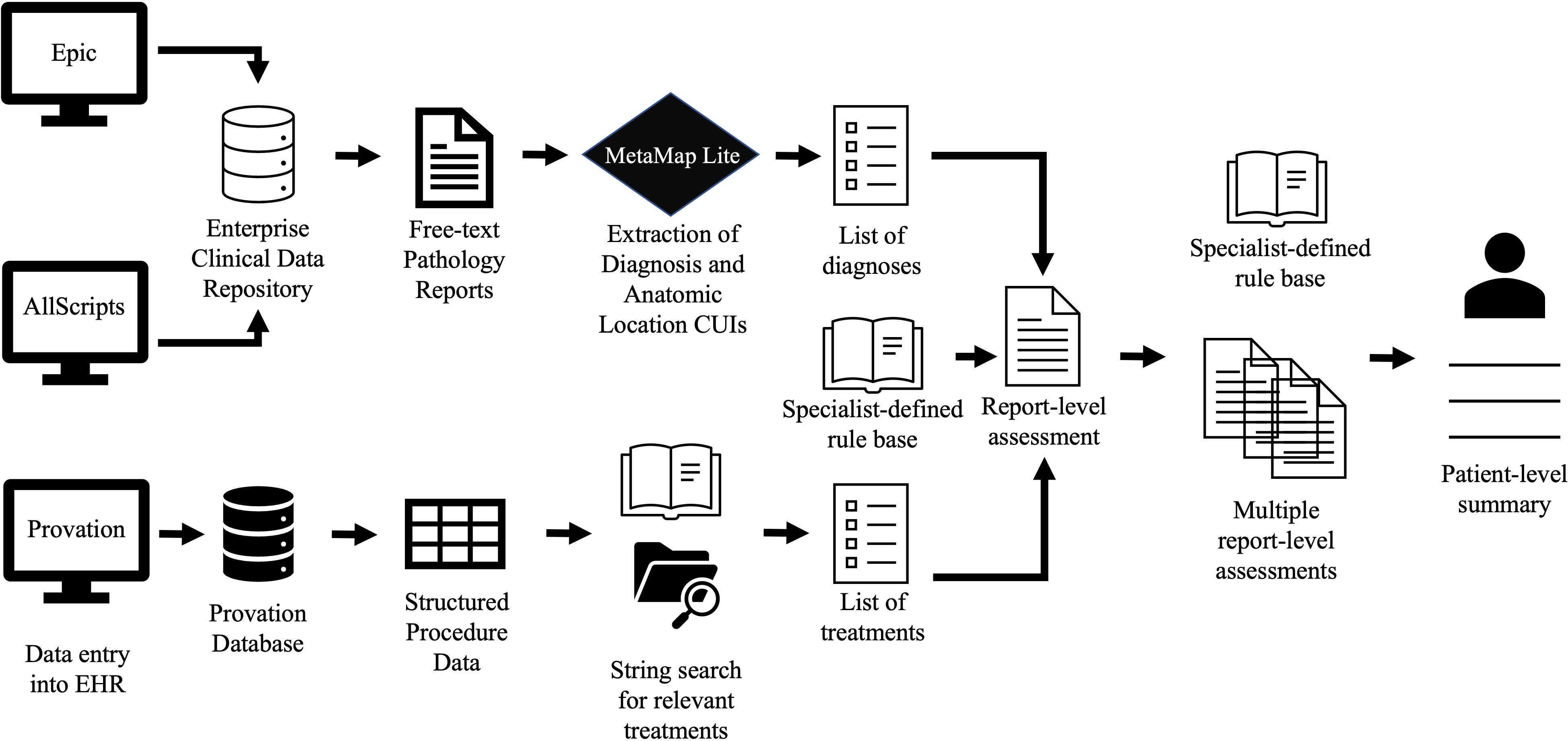
Summary of system architecture. Free text upper endoscopy pathology reports from the current and historic EHR systems are sourced from the CUIMC Clinical Data Repository (CDR). Concept Unique Identifiers (CUIs) pertaining to diagnoses are extracted from pathology reports using MetaMap Lite. Structured procedure-related data, including endotherapies, are extracted from the ProVation endoscopy documentation database and merged with the corresponding pathology report diagnoses to create report-level assessments. A patient-level summary is then generated from multiple report-level assessments according to the clinical logic specified in a rule base.

We applied MetaMapLite to the processed pathology text, extracting Unified Medical Language System (UMLS) concept unique identifiers (CUIs) that represent medical concepts. We filtered the CUIs to include esophageal or gastroesophageal anatomic concepts and BE-related histological diagnosis concepts. Specimen-level diagnoses were derived by linking sequential filtered anatomic location and histologic diagnosis concepts within the pathology report text. The worst specimen-level BE diagnosis concept within a given pathology report defined the procedure-level diagnosis concept. A final set of rules reduced the MetaMapLite-generated diagnosis concepts to the previously defined full set of BE diagnosis labels (*no BE*, *no dysplasia*, *indefinite for dysplasia*, *LGD*, *HGD*, and *EAC*. Simplified binary diagnosis labels for dysplasia and EAC were derived from the full set of labels. BE-related endotherapies were extracted from structured endoscopy impression and maneuver endoscopy report data using string searches. Each endoscopy could have multiple endotherapy classifications.

### Calculation of Patient-level Quality Metrics

We applied the NLP pipeline to the original cohort of patients who had upper endoscopy for BE-related indications, with the goal of identifying all patients who initiated endoscopic eradication therapy (EET) for BE during the period of interest. Manual review to exclude those not meeting the clinical definition of BE was not performed as it was assumed that patients with a BE-related indication for endoscopy and evidence of endotherapy had a confirmed diagnosis of BE. An additional rule-based algorithm was applied to the resulting procedure-level data to identify the dates of key clinical events including endotherapy initiation, ongoing endotherapy, CE-D (complete eradication of dysplasia), and CE-IM (complete eradication of intestinal metaplasia). We defined EET initiation as the date of the first resection of visible abnormalities (EMR or ESD) or ablation (RFA or cryotherapy) where there was a concurrent or immediately preceding histologic diagnosis of BE-related dysplasia. APC was not considered to be a valid initial EET modality as it was primarily used as a touch-up treatment. Ongoing endotherapy was defined as the inclusive period between the endotherapy initiation and CE-IM. CE-IM was defined as the first date on which there was no histologic evidence of BE or BE-related neoplasias and no documented endotherapy in a patient undergoing EET. CE-D was defined as the first date on which there was histologic evidence of dysplasia and no documented endotherapy in a patient undergoing EET. All patients who had an NLP-derived EET initiation date, were between the ages of 40 and 80, and did not undergo esophagectomy (past or future) were included in the EET set. Ground truth labels for key clinical event dates were assigned via manual review of all available EHR data. Using the NLP-derived key clinical event dates and additional algorithmic rules, we determined patient-level variables such as worst pre-EET diagnosis, endotherapy modalities received, endotherapy duration, time to CE-IM, and time to CE-D.

### Statistical Analysis

For the validation set, Kaplan’s Kappa was calculated between the 2 gastroenterologist annotators to determine interrater reliability. Performance metrics comparing the NLP tool to the ground truth labels were calculated for all datasets. Macro accuracy, precision (positive predictive value), recall (sensitivity), and F1-score (the harmonic mean of precision and recall) were determined for the multiclass NLP classifier at the global level, as well as for diagnosis and endotherapy alone. Performance metrics for the binary diagnosis classifiers were determined at the diagnosis level only. Discrepancies between the ground truth and NLP labels were identified and qualitatively grouped by presumed error etiology.

## RESULTS

### Dataset Characteristics

977 patients underwent BE surveillance or endotherapy during the period of interest. After applying exclusion criteria, the development and validation sets included 99 and 115 patients respectively (**Figure 1**). Out of the 377 endoscopies in the development set, 43.5% found a diagnosis of BE or worse, 15.9% found dysplasia or worse, and 2.9% found adenocarcinoma (**Table 1**). There was a similar histologic diagnosis distribution in the validation set (44.9%, 12.5%, 5.5% out of 400 endoscopies). In both the development and validation sets, similar proportions of the endoscopies had associated endotherapy data (29.4% versus 29.3%). The distribution of endotherapy was also similar between the two sets, except that the validation set had a higher proportion of radiofrequency ablation (15.0% vs. 11.4%) and a lower proportion of cryotherapy (2.0% vs. 4.5%) compared to the development set.

**Table 1:** Ground-Truth Characteristics of the Development and Validation Datasets.

|  | Development Set<br>(n = 99 patients)<br>(n = 377 procedures) | Validation Set<br>(n = 115 patients)<br>(n = 399 procedures) |
| --- | --- | --- |
| **Patient Characteristics** |  |  |
| Median procedures per patient (IQR) | 3.0 (1.5-5.0) | 3.0 (1.0-5.0) |
| Median endoscopy reports per patient (IQR) | 3.0 (1.0-5.0) | 2.0 (1.0-4.0) |
| Median pathology reports per patient (IQR) | 2.0 (1.0-4.0) | 2.0 (1.0-4.0) |
| **Procedure-level BE-related diagnosis, n (%)** |  |  |
| No specimens | 74 (19.6%) | 79 (19.8%) |
| No evidence of BE | 139 (36.9%) | 141 (35.3%) |
| BE with no dysplasia | 89 (23.6%) | 101 (25.3%) |
| BE, indefinite for dysplasia | 15 (4.0%) | 22 (5.5%) |
| BE with low-grade dysplasia | 18 (4.8%) | 8 (2.0%) |
| BE with high-grade dysplasia | 31 (8.2%) | 26 (6.5%) |
| Esophageal adenocarcinoma | 11 (2.9%) | 22 (5.5%) |
| **Procedure-level BE-related endotherapy,\* n (%)** |  |  |
| No endotherapy | 266 (70.6%) | 282 (70.7%) |
| Endoscopic mucosal resection | 29 (7.7%) | 28 (7.0%) |
| Endoscopic submucosal dissection | 1 (0.3%) | 1 (0.3%) |
| Radiofrequency ablation | 43 (11.4%) | 60 (15.0%) |
| Argon plasma coagulation | 35 (9.3%) | 32 (8.0%) |
| Cryotherapy | 17 (4.5%) | 8 (2.0%) |

BE: Barrett's Esophagus
\* More than one endotherapy can be performed per procedure

### Clinical Data Labelling Performance

Global (both diagnosis and endotherapy) recall, precision, accuracy, and F1-score for the multiclass classifier were 0.976, 0.970, 0.985, and 0.972 respectively in the validation set (**Table 2**). The binary dysplasia classifier (recall: 1.000, precision: 0.966, accuracy: 0.990, F1-score: 0.982) had improved performance across all metrics compared to the multiclass classifier (diagnosis-only recall: 0.973, precision: 0.946, accuracy: 0.975, F1-score: 0.958). Unsurprisingly, there was a drop in multiclass classifier performance in validation set compared to the development set. However, binary classifier performance was maintained in the validation set.

**Table 2:** Barrett’s Esophagus Endoscopic Outcome Classifier Performance Metrics

| **Multiclass Classifier (all labels)** |  |  |  |  |
| --- | --- | --- | --- | --- |
|  | **Recall** | **Precision** | **Accuracy** | **F1-Score** |
| **Development Set** |  |  |  |  |
| Global Performance | 0.985 | 0.989 | 0.995 | 0.986 |
| Diagnosis-only Performance | 0.970 | 0.977 | 0.989 | 0.971 |
| Endotherapy-only Performance | 1.000 | 1.000 | 1.000 | 1.000 |
| **Validation Set** |  |  |  |  |
| Global Performance | 0.976 | 0.970 | 0.985 | 0.972 |
| Diagnosis-only Performance | 0.973 | 0.946 | 0.975 | 0.958 |
| Endotherapy-only Performance | 0.979 | 0.995 | 0.995 | 0.986 |
| **Binary Dysplasia Classifier (Presence/absence of Dysplasia or Worse)** |  |  |  |  |
|  | **Recall** | **Precision** | **Accuracy** | **F1-Score** |
| **Development Set** |  |  |  |  |
| Diagnosis-only Performance | 1.000 | 0.938 | 0.989 | 0.968 |
| **Validation Set** |  |  |  |  |
| Diagnosis-only Performance | 1.000 | 0.966 | 0.990 | 0.982 |

| Binary EAC Classifier (Presence/absence of Esophageal Adenocarcinoma) |  |  |  |  |
| --- | --- | --- | --- | --- |
|  | Recall | Precision | Accuracy | F1-Score |
| **Development Set**<br>Diagnosis-only Performance | 1.000 | 1.000 | 1.000 | 1.000 |
| **Validation Set**<br>Diagnosis-only Performance | 1.000 | 1.000 | 0.995 | 0.990 |

### Development and Validation Set Classifier Error Analysis

Diagnosis classification errors in the development set occurred due to diagnostic uncertainty narratives (n=3) and the mention of a prior diagnosis within the addendum text (n=1). A representative diagnostic uncertainty narrative is: *“…opinions varied from reactive to low-grade dysplasia. In my opinion, indefinite is the best classification*”. In this example, the classifier selected the most severe diagnosis without negation. It cannot process a freeform declarative statement like “*In my opinion, indefinite is the best classification.*” The presence of a prior diagnosis in the addendum text similarly cannot be resolved by our algorithm, as our pathology text NLP algorithm cannot understand temporal context in a more granular way than excluding note sub-sections that generally contain prior diagnoses (ie. The *Clinical History* sub-section).

Diagnosis classification errors due to diagnostic uncertainty (n=5) similar to those observed in the development set were also present in the validation set. Two additional categories of errors emerged in the validation set: novel text patterns that did not fit into the existing rules (n=4) and missing pathology report data (n=1). Novel text patterns that were not captured by the NLP system included a missing space (“*Barrett’sesophagus*”), a novel synonym for intestinal metaplasia (“*rare goblet cells*”), a new anatomic location pattern (“*Barrett’s patchy*”), and the presence of a publication title cited as a reference in the pathology report containing a more severe diagnosis than the remainder of the pathology report text (“*Eosinophilic infiltration of the esophagus following endoscopic ablation of Barrett’s neoplasia*”)

Endotherapy classification errors were related to the non-therapeutic use of ablation (n=2). In one case, cryotherapy was attempted, but aborted due to patient instability. In the other, APC was used only for marking lesion boundaries, rather than ablating abnormal tissue.

### BE Quality Metric Assessment

The EET dataset included 77 patients whose upper endoscopy history during the period of interest comprised of 554 endoscopies, of which 254 (45.8%) involved endotherapy and 384 (69.3%) had associated pathology reports (dysplasia: n=133) (**Table 3)**. Within this dataset, there were 12 patients and 101 endoscopies from the development set and 20 patients and 147 endoscopies from the validation set. Multiclass classifier performance slightly deteriorated in the EET set (global recall: 0.963, precision: 0.981, accuracy: 0.988, and F1-score: 0.970) compared to the validation set. Classification errors at this level were related to diagnostic uncertainty (n=7), endotherapy misclassification (n=2), and BE diagnosis misclassification (n=3). Notably, no novel text patterns were observed in this additional dataset. New BE diagnosis misclassification errors in the quality metric NLP process were attributed to the lack of a process for excluding patients who did not have a clinical diagnosis of BE. Patients who did not have BE underwent endotherapy for gastric cancer (n=2) and esophageal dysplasia (n=1) and also had endoscopic procedures for BE-related indications.

**Table 3:** Characteristics of Patients Undergoing Endoscopic Eradication Therapy for Barrett’s Esophagus (n = 77 patients, n = 554 procedures)

| Patient Characteristics |  |
| --- | --- |
| Age, avg +/- std | 67.5 +/- 8.8 |
| Sex, n (%) |  |
| Male | 64 (83.1%) |
| Female | 13 (16.9%) |
| Race, n (%) |  |
| White | 64 (83.1%) |
| Non-White | 4 (5.2%) |
| Other/Unknown/Declined | 9 (11.7%) |
| Ethnicity, n (%) |  |
| Not Hispanic | 57 (74.0%) |
| Hispanic | 2 (2.6%) |
| Unknown/Declined | 18 (23.4%) |
| Median procedures per patient (IQR) | 7.0 (5.0-8.0) |
| Median endoscopy reports per patient (IQR) | 7.0 (5.0-8.0) |
| Median pathology reports per patient (IQR) | 5.0 (3.0-7.0) |
| **Procedure-level BE-related diagnosis, n (%)** |  |
| No specimens | 171 (30.9%) |
| No evidence of BE | 125 (22.6%) |
| BE with no dysplasia | 66 (11.9%) |
| BE, indefinite for dysplasia | 27 (4.9%) |
| BE with low-grade dysplasia | 47 (8.5%) |
| BE with high-grade dysplasia | 91 (16.4%) |
| Esophageal adenocarcinoma | 27 (4.9%) |
| **Procedure-level BE-related endotherapy,\* n (%)** |  |
| No endotherapy | 300 (54.2%) |
| Endoscopic mucosal resection | 121 (21.8%) |
| Endoscopic submucosal dissection | 3 (0.5%) |
| Radiofrequency ablation | 230 (41.5%) |
| Argon plasma coagulation | 86 (15.5%) |
| Cryotherapy | 29 (5.2%) |

BE: Barrett's Esophagus
\* More than one endotherapy can be performed per procedure

Despite the diagnosis and endotherapy labelling errors, accuracy for the key clinical event labels was uniformly high: EET start (97.4%), CE-D (100.0%), and CE-IM (100.0%). Using the combined diagnosis, endotherapy, and event labels, our BE quality metric algorithm successfully automated extraction of key patient-level measures: pretherapy diagnosis (LGD: 20.8%, HGD: 58.4%, EAC: 20.8%), endotherapy time (median: 8.3 months), time to CED (median: 9.1 months), and time to CE-IM (median: 11.1 months) (**Table 4**).

**Table 4:**
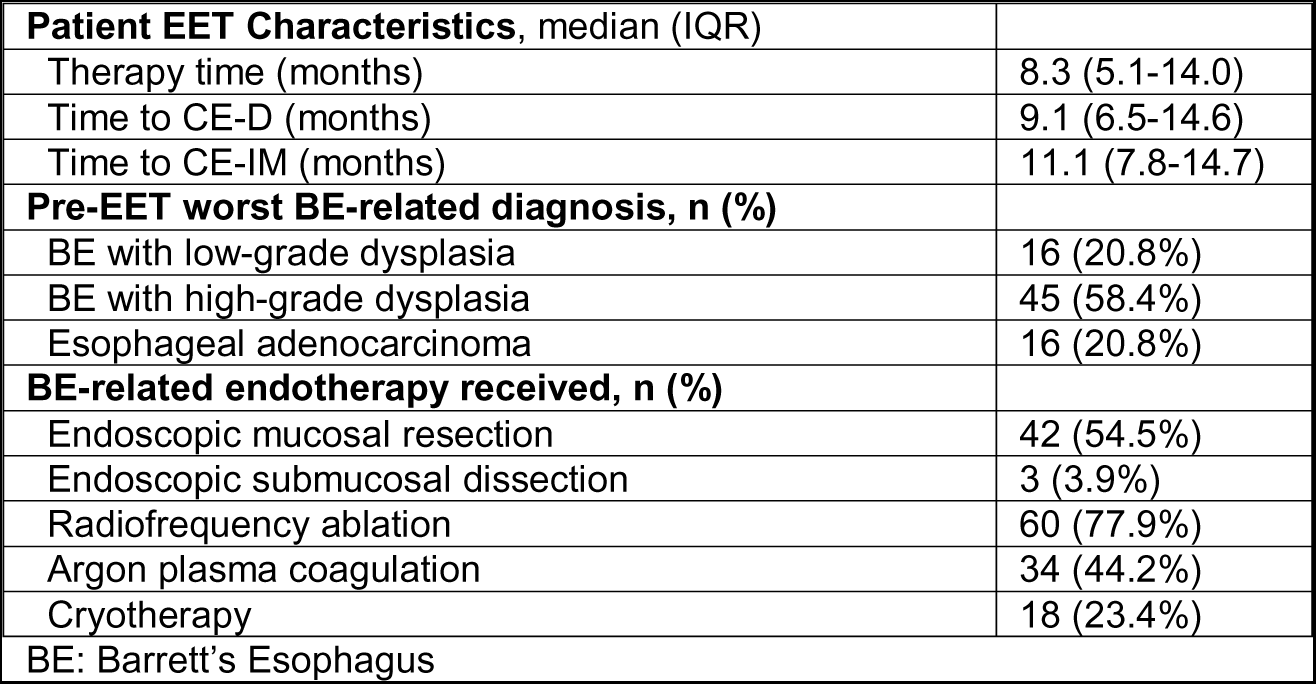
Patient-level Data for Patients Undergoing Endoscopic Eradication Therapy for Barrett’s Esophagus (n = 77 patients)

## DISCUSSION

We have developed a novel NLP pipeline for automated extraction of key endoscopic BE surveillance and treatment data. The datasets generated by the pipeline additionally facilitate expedited manual data review for additional metrics not directly obtained by the pipeline. The pipeline performed well both when using all relevant BE diagnoses as well as simplified binary diagnoses (presence/absence of dysplasia or cancer). We reliably extracted key clinical events and higher-level patient-level variables such as worst pre-EET histologic diagnosis, endotherapy time, and time to CE-IM. This represents a significant improvement over current labor-intensive data extraction approaches using manual chart review.

There is a single prior study of NLP for BE clinical data extraction, in which a machine-learning-based approach was used to extract dysplasia diagnoses from the pathology reports of randomly sampled patients with BE. This approach yielded 0.987 accuracy, 0.923 recall, 1.000 precision, and an F1-score of 0.960 in a validation set with a similar number of dysplasia cases (29). For the binary BE dysplasia classification task, our method outperformed the prior method across all measures except precision (positive predictive value). While the prior study was validated in a national database, our NLP pipeline provides higher granularity BE-related histology data as well as additional clinical data that allows the extraction of higher-level measures like BE endoscopy quality metrics.

Our rule-based algorithm built on the open-source MetaMapLite NLP tool enables algorithmic transparency, or the ability to understand model decision-making. When interrogating our pipeline, we found it had difficulty parsing novel text patterns like misspellings and the complex, unstructured narratives used to express diagnostic uncertainty. Diagnostic uncertainty is an especially common issue in BE pathology report text, as BE-related dysplasia diagnosis has poor interobserver agreement (31). While understandable and computationally efficient, our rule-based NLP approach hampers the generalizability of our system. The data pre-processing methods and rules based on text patterns would need to be validated before use with another EHR system or even with different time periods in the same EHR.

In the future, deep learning approaches could allow a more generalizable means of extracting BE pathology diagnoses from free text notes thereby reducing the need to develop complex rule-based algorithms (32–34). However, even this approach has limitations, as privacy concerns limit the transportability of model weights across institutions and deep learning models can still be prone to over-fitting to the development dataset. Novel large language models like LLaMA, Med-PALM2, and GPT-4 hold the promise of facilitating the development of NLP pipelines for clinical text with less text preprocessing and no development dataset or very small development datasets (35–38). With the time saved using such methods, future iterations of this and related systems could incorporate additional metrics relevant to BE quality, such as adherence to the Seattle protocol, use of appropriate surveillance intervals, and use of emerging risk stratification biomarkers such as p53 (8, 39).

## CONCLUSION

We have developed and internally validated an automated NLP pipeline that extracts the full range of BE-related histological diagnoses, BE-related endoscopic therapies, and key BE-related quality metrics using both pathology reports and structured endoscopy report data. Future research is needed to extend this approach to novel large language model technologies and to assess generalizability to other institutions.

### Ethics Statement

The Institutional Review Board of Columbia University Irving Medical Center reviewed the protocol for this study and gave ethical approval for this work.

### Data Availability Statement

All data produced in the present study are available upon reasonable request to the authors.

### Financial Support

This study was supported by grants from the NIH/NCI (P30CA013696, R01CA238433), NIH/NCATS (UL1TR001873), NIH/NLM (T15LM007079, R01LM009886), and NIH/NIGMS (GM131905)

## Data Availability

All data produced in the present study are available upon reasonable request to the authors.

