## Supplementary Tables for "Natural Language Processing Can Automate Extraction of Barrett’s Esophagus Endoscopy Quality Metrics"

**Supplementary Table 1.** Confusion Matrix for Procedure-Level Diagnoses, Development Set

| **True Labels** | **Predicted Labels** | | | | | | |
| --- | --- | --- | --- | --- | --- | --- | --- |
|  | **No Label** | **No BE** | **NDBE** | **Indefinite** | **LGD** | **HGD** | **EAC** |
| **No Label** | 74 | 0 | 0 | 0 | 0 | 0 | 0 |
| **No BE** | 0 | 139 | 0 | 0 | 0 | 0 | 0 |
| **NDBE** | 0 | 0 | 88 | 0 | 0 | 1 | 0 |
| **Indefinite** | 0 | 0 | 0 | 12 | 2 | 1 | 0 |
| **LGD** | 0 | 0 | 0 | 0 | 18 | 0 | 0 |
| **HGD** | 0 | 0 | 0 | 0 | 0 | 31 | 0 |
| **EAC** | 0 | 0 | 0 | 0 | 0 | 0 | 11 |

**BE**: Barrett’s esophagus; **NDBE**: non-dysplastic Barrett’s esophagus; **Indefinite**: indefinite for dysplasia; **LGD**: low-grade dysplasia; **HGD**: high-grade dysplasia; **EAC**: esophageal adenocarcinoma.

**Supplementary Table 2.** Confusion Matrix for Procedure-Level Diagnoses, Validation Set

| **True Labels** | **Predicted Labels** | | | | | | |
| --- | --- | --- | --- | --- | --- | --- | --- |
|  | **No Label** | **No BE** | **NDBE** | **Indefinite** | **LGD** | **HGD** | **EAC** |
| **No Label** | 79 | 0 | 0 | 0 | 0 | 0 | 0 |
| **No BE** | 0 | 138 | 1 | 2 | 0 | 0 | 0 |
| **NDBE** | 1 | 3 | 97 | 0 | 0 | 0 | 0 |
| **Indefinite** | 0 | 0 | 0 | 20 | 2 | 0 | 0 |
| **LGD** | 0 | 0 | 0 | 0 | 8 | 0 | 0 |
| **HGD** | 0 | 0 | 0 | 0 | 0 | 25 | 1 |
| **EAC** | 0 | 0 | 0 | 0 | 0 | 0 | 22 |

**BE**: Barrett’s esophagus; **NDBE**: non-dysplastic Barrett’s esophagus; **Indefinite**: indefinite for dysplasia; **LGD**: low-grade dysplasia; **HGD**: high-grade dysplasia; **EAC**: esophageal adenocarcinoma.
